# The neural basis of frontotemporal dementia (FTD): insights from ALE meta-analyses of four FTD subtypes encompassing 8,057 patients

**DOI:** 10.1101/2025.06.02.25328809

**Authors:** Zlatomira G. Ilchovska, Justine Lockwood, Elizabeth Proctor, Akram Hosseini, Matthew A. Lambon Ralph, JeYoung Jung

**Affiliations:** School of Psychology, University of Nottingham, Nottingham, UK; Division of Clinical Neuroscience, University of Nottingham, Nottingham, UK; Department of Neurology, Nottingham University Hospitals NHS Trust, Queen’s Medical Centre, Nottingham, Nottingham, UK; Centre for Dementia, Institute of Mental Health, University of Nottingham, UK; MRC Cognition and Brain Sciences Unit, University of Cambridge, Cambridge, UK; NIHR Biomedical Research Centre, University of Nottingham, UK

**Author notes:** Corresponding author: Dr JeYoung Jung, School of Psychology, University of Nottingham, University Park, Nottingham NG7 2RD UK.

**Keywords:** frontotemporal dementia (FTD), primary progressive aphasia (PPA), behavioural variant FTD, nonfluent variant PPA, semantic dementia (SD), logopenic variant PPA (lvPPA), meta-analysis

## Abstract

Frontotemporal dementia (FTD) is a complex neurodegenerative syndrome primarily affecting the frontal and temporal lobes, resulting in deficits in behaviour, executive function, and language. As the leading cause of early-onset dementia, FTD includes several subtypes, notably behavioural variant FTD (bvFTD) and primary progressive aphasia (PPA). PPA is further subdivided into three main variants: non-fluent variant PPA (nfvPPA), semantic variant PPA (svPPA)/semantic dementia (SD), and logopenic variant PPA (lvPPA), each characterised by distinct language and cognitive impairments. However, despite these clinical distinctions, increasing evidence suggests overlapping behavioural symptoms and neural abnormalities across FTD syndromes, challenging the traditional view of them as mutually exclusive. To explore the neural basis of FTD, we conducted an activation likelihood estimation meta-analysis, synthesising data from structural and functional neuroimaging studies across four major FTD subtypes. Our analysis included coordinate-based data from 114 studies comprising 8,057 FTD patients. Results revealed widespread brain abnormalities in FTD compared to healthy controls, encompassing the frontal, temporal, and parietal lobes, as well as subcortical and limbic regions such as the basal ganglia and thalamus. This widespread degeneration aligns with the broad spectrum of cognitive and behavioural symptoms seen in FTD. Each FTD subtype demonstrated distinct, yet partially overlapping, patterns of degeneration. BvFTD showed prominent degeneration in the frontal and medial temporal lobes, insula, cingulate cortex, and limbic system, consistent with impairments in social cognition and disinhibition. SvPPA/SD exhibited focal atrophy in the anterior temporal lobes, disrupting the semantic network and impairing semantic processing. NfvPPA was associated with degeneration in the speech production network, particularly the insula and inferior frontal gyrus. LvPPA displayed left-lateralised abnormalities in the posterior temporal and inferior parietal lobes, affecting language function. Additionally, FTD subtypes exhibited overlapping patterns of degeneration, contributing to shared cognitive and language deficits. For example, svPPA/SD and bvFTD both showed degeneration in the anterior temporal lobe and amygdala, linked to socio-emotional impairments. BvFTD and nfvPPA shared degeneration in the left frontal cortex and insula, suggesting common executive dysfunction. Similarly, lvPPA and svPPA/SD both showed abnormalities in the left lateral temporal lobe, linking these regions to semantic cognition deficits observed in both variants. These findings highlight the mixture of clinical heterogeneity of each FTD subtype as well as their overlapping features. This study offers new insights into the neural bases of recent proposals that FTD spans a transdiagnostic multidimensional phenotype-geometry and may aid in refining diagnostic criteria and developing targeted interventions for this group of neurodegenerative disorders.

## 1. Introduction

Frontotemporal dementia (FTD) is a neurodegenerative syndrome characterized by the progressive deterioration of the frontal and temporal lobes, resulting in deficits in behaviour, executive function and language ^1–3^. This is the most prevalent type of early-onset dementia and ranks the third most common form of dementia overall, following Alzheimer’s disease and Lewy body disease ^4^. FTD encompasses various clinical syndromes distinguished by predominant features. Approximately half of cases manifest as the behavioural variant FTD (bvFTD), characterized by disruptions in social behaviour and personality ^5^, while the remainder fall under the label of primary progressive aphasia (PPA), demonstrating language decline ^6^. PPA includes three main variants: non-fluent variant PPA (nfvPPA) characterized by speech production difficulties ^7^, semantic variant PPA (svPPA) or semantic dementia (SD) exhibiting multimodal semantic memory degradation ^8^, and logopenic variant PPA (lvPPA) marked by impairments in word finding, sentence repetition and syntactic comprehension ^9^. Although the paradigmatic exemplar cases are relatively distinct from one another, growing research has shown that there are important variations with each FTD subtype, clear graded overlaps between subtypes, a substantial number of patients who do not fall into any one category ^10–16^. The considerable clinical and pathological heterogeneity of FTD adds complexity to its recognition and diagnosis ^2,17^. While recent diagnostic criteria reflect this heterogeneity ^5^, there may be improvements in early detection and diagnosis by incorporating neuroimaging ^18^. Here, we aim to provide an overview of the neural basis of FTD – bvFTD and the three PPA variants, employing coordinate-based meta-analysis (CBMA) to identify both distinct and overlapping patterns of brain degeneration across subtypes.

Neuroimaging studies have contributed to advancing our understanding of FTD for a review, see ^18^. Studies utilizing structural magnetic resonance imaging and fluorodeoxyglucose positron emission tomography (FGD-PET) have revealed distinct patterns of brain atrophy and hypometabolism corresponding to the clinical and pathological variants of FTD. Diffusion tensor imaging (DTI) and resting-state functional magnetic resonance imaging (rsfMRI) have delineated structural and functional changes in brain connectivity associated with FTD. In bvFTD, brain atrophy impacts various regions including the orbitofrontal cortex (OFC), medial/lateral prefrontal cortex, anterior cingulate cortex (ACC), insula, and subcortical areas ^19–21^. White matter damage involves pathways, including the superior longitudinal fasciculus and corpus callosum ^22,23^. Previous studies of BvFTD have found decreased functional connectivity within the salience network, including the prefrontal cortex, insula, and ACC ^24–26^. SvPPA/SD is linked to atrophy and hypometabolism in the anterior temporal lobe (ATL) ^8,27,28^ along with damage to the uncinate fasciculus and the inferior longitudinal fasciculus ^29^. NfvPPA involves atrophy of the left inferior frontal gyrus (IFG), insula and premotor cortex ^30–32^, while lvPPA is associated with atrophy in the left superior temporal and inferior parietal lobe (IPL) ^33,34^. These structural and functional alterations are associated with the clinical symptoms of the subtypes of FTD.

Individual neuroimaging studies in FTD often yield inconsistent findings due to various factors, including small cohort sizes, diverse clinical samples, different imaging techniques, and methodological variability. In contrast, meta-analysis addresses these challenges by aggregating data from multiple studies, providing more comprehensive and robust findings. Meta-analysis can unveil patterns or trends across studies, deliver more precise estimates of effect sizes, and enhance statistical power by pooling data from various sources. Through synthesizing findings from multiple studies, meta-analysis enhances the reliability and validity of conclusions drawn from neuroimaging research ^35^. Recent works utilizing CBMA have shown consensus on brain alterations in certain FTD subtypes. For example, Kamalian and colleagues ^36^ reported convergent abnormalities in the salience network and subcortical regions in bvFTD. Similarly, Yang and colleagues ^37^ conducted a meta-analysis on voxel-based morphometry (VBM) studies and demonstrated the common atrophy in the bilateral ATL and caudate in svPPA/SD. However, meta-analysis in FTD is typically limited to a specific type of FTD and no study has provided a comprehensive overview of brain alterations across FTD spectrum, encompassing both bvFTD and PPA.

A growing body of research has highlighted substantial variability within each FTD subtype, with graded overlaps across subtypes and a considerable proportion of patients presenting with mixed features that do not fit neatly into a single diagnostic category ^11–16^. The emerging picture is that, rather than mutually-exclusive diagnostic category boundaries, there is a systematic transdiagnostic multidimensional with patient varying along one or more cognitive-behavioural-language dimensions ^11,15,16,38,39^ – with the classical exemplar cases sitting in distinct positions of this multidimensional space. For example, bvFTD and SD/svPPA form a clear frontotemporal continuum associated with behavioural and executive impairment as atrophy affects frontal regions and multimodal semantic impairment resulting from ATL pathology ^13,16,38,39^. Likewise there can be overlap between bvFTD and some subcortical dementias in motor features and executive-behavioural changes such as apathy ^1,2,11^, and when language features appear in some subcortical disorders then it mirrors the nfvPPA phenotype ^40^. Given these partially overlapping, graded differences between the FTD disorder, researchers have hypothesised that each patient’s exact phenotype reflects the varying distribution of atrophy ^14,41,42^. Further supporting it, a recent systematic review showed that while bvFTD is primarily characterised by socio-emotional and behavioural changes, many patients also exhibit extensive but heterogeneous speech and language impairments, including lexico-semantic, orthographic, and prosodic deficits, which overlap with those seen in PPA ^43^. Conversely, socio-emotional deficits such as impaired emotion recognition, empathy, and theory of mind are frequently reported across PPA variants ^44–46^, especially in svPPA/SD, where they appear similar to those in bvFTD, reflecting the shared frontotemporal changes ^13,16,38^. In response to this complexity, a transdiagnostic approach has emerged in FTD to better understand its aetiology and pathophysiology and develop more effective therapies. Murley et al. ^11^ examined 310 patients with FTD spectrum syndromes, finding overlapping clinical features across diagnostic groups. Sixty-two percent of patients exhibited features consistent with more than one syndrome. Patterns of brain atrophy were found to covary across diagnostic groups, revealing brain-behaviour relationships that form a continuous spectrum rather than discrete diagnostic categories. These findings challenge the notion of FTD syndromes as mutually exclusive categories defined solely by clinical features or brain structure changes. Instead, they suggest a multidimensional phenotypical geometry wherein FTD syndromes exist as overlapping clusters within a broader cognitive-neuroanatomical landscape ^11,13,14,16,38,39^. Here, we aim to establish a convergent understanding of neural basis across the FTD spectrum.

The main goal of this study was to elucidate the neural basis of FTD spectrum through a CBMA of published neuroimaging studies. We assessed the consensus of structural and functional alterations in four subtypes of FTD – bvFTD, svPPA/SD, nfvPPA and lvPPA, employing activation likelihood estimation (ALE) meta-analysis. Specifically, we examined (i) the overall convergence of findings from structural and functional neuroimaging studies, (ii) the concurrence within each FTD variant, and (iii) the overlaps and distinctiveness between the different FTD variants. Through this examination, we aimed to determine whether structural and functional alterations can be linked to clinical phenotypes in FTD, contributing to both its heterogeneity and shared characteristics. Our results would provide insights into discrepancies in the existing literature on FTD.

## 2. Methods and materials

This large-scale CBMA was conducted in accordance with the latest best-practice guidelines for neuroimaging meta-analyses ^35^ and followed the PRISMA (Preferred Reporting Items for Systematic Reviews and Meta-Analyses) statement ^47^. The study protocol was pre-registered on the International Prospective Register of Systematic Reviews (PROSPERO) under the code CRD42023449002.

### 2.1 Search Strategy and Criteria

We conducted a PubMed and Google Scholar search on articles published up to Oct 2023 on the four selected types of FTD (bvFTD, svFTD/SD, nfvPPA and lvPPA). Our search strategy involved the following combinations of keywords: (behavioural variant frontotemporal dementia OR bvFTD OR frontal variant frontotemporal dementia OR frontotemporal dementia OR Pick’s disease OR primary progressive aphasia OR semantic dementia OR semantic variant PPA OR svPPA OR progressive non fluent aphasia OR agrammatic variant PPA OR nonfluent variant PPA OR nfvPPA OR logopenic primary progressive aphasia phonological variant PPA OR logopenic variant PPA OR lvPPA) AND (functional magnetic resonance imaging OR fMRI OR voxel-based morphometry OR VBM OR positron emission tomography OR PET).

We searched for articles that met the following criteria: 1) published in English; 2) contained a between-group comparison of a healthy control (HC) group and clinically diagnosed FTD patients, where the patient group has no concurrent psychiatric disorder, other forms of dementia, neurological symptoms or any history of substance abuse; 3) included a minimum of 6 participants per group; 4) employed task-based fMRI, resting-state fMRI, VBM or FDG-PET neuroimaging techniques for data collection; 5) performed a whole-brain analysis; 6) reported brain coordinates in Talairach or MNI standard space; 7) for structural imaging studies, reported grey matter coordinates (excluding white matter coordinates). We excluded studies using seed-based functional connectivity analysis, DTI, and cortical thickness measurements. After removing duplicates, three authors (Z.I, J.L. & E.P.) independently reviewed the unique records, initially by screening abstracts and subsequently by reading the full texts of potentially relevant articles. Any disagreements among the three raters were resolved by an additional reviewer (J.J.). Figure 1 illustrates a flow diagram based on the PRISMA guidelines, illustrating the inclusion and exclusion criteria applied during the study selection process.

**Figure 1.**
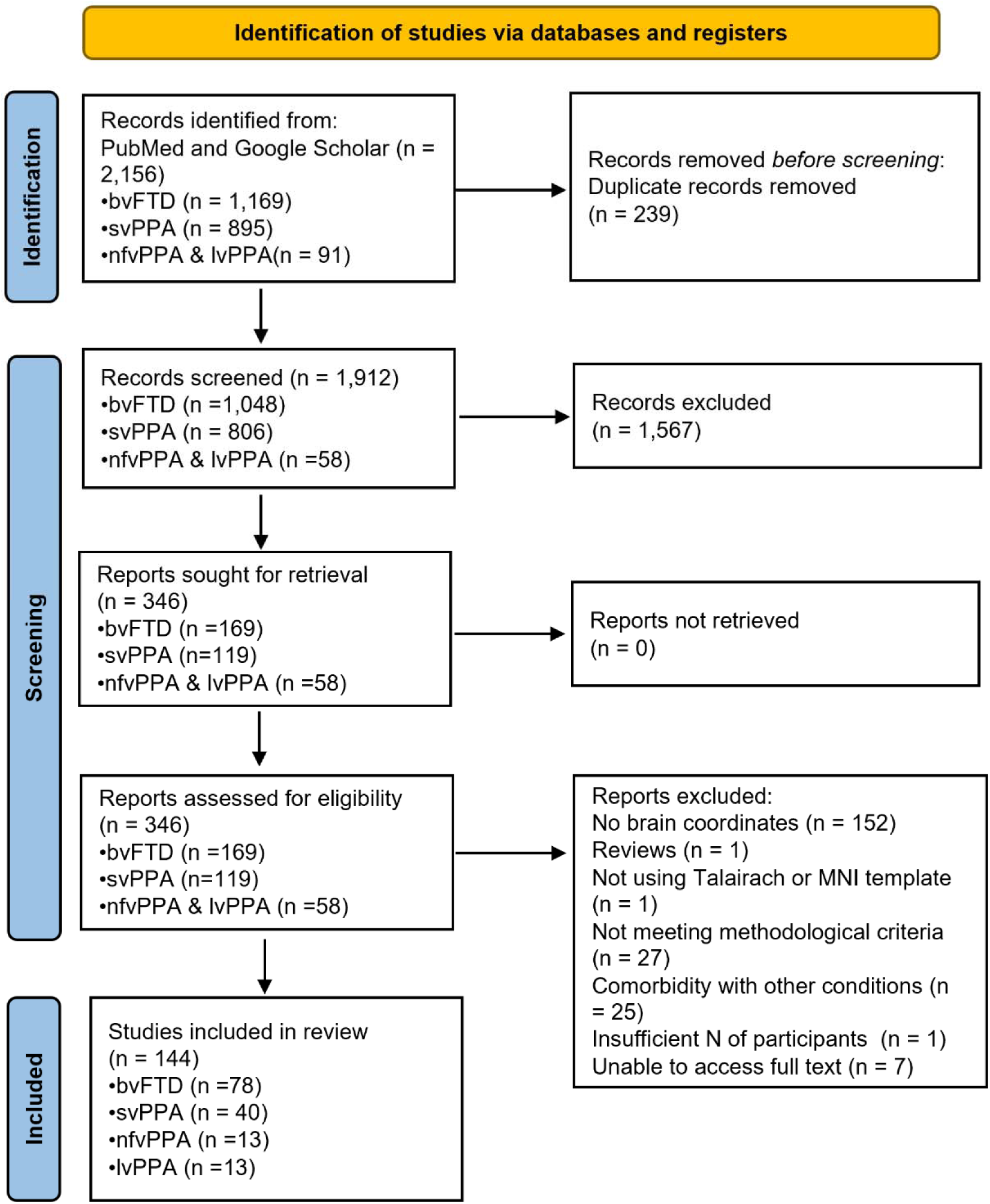
PRISMA flow diagram of the study selection process.

### 2.2. Activation likelihood estimation analysis

The updated ALE algorithm using Ginger ALE (version 3.0.2) was employed to detect convergent patterns of brain alterations, demonstrating that the convergence of reported coordinates across experiments exceeds what would be expected from random spatial associations ^48^. This method tests the spatial convergence of reported coordinates for differences between FTD patients and healthy controls (HCs) against the null hypothesis of randomly distributed findings across the brain. Each experiment’s foci were modelled as 3D Gaussian kernels to represent the uncertainty about the peak coordinates’ locations. A modelled activation map was then created for each experiment by combining all the convolved foci. The union of these maps for all included experiments resulted in an ALE score map, representing the voxel-wise likelihood of convergent findings at each brain location. To distinguish true convergence from random overlap, the ALE score map was statistically tested against a null distribution of randomly distributed findings, with a p<0.05 cluster-level family-wise error (cFWE) correction for multiple comparisons (a cluster-forming p < 0.001 with thresholding 1,000 permutations). Prior to the analysis, all Talairach coordinates were transformed into MNI space using the transform called icbm2tal developed by Laird et al ^49^.

To test the spatial convergence of reported coordinates for differences between patients and HCs, we performed separate ALE meta-analyses for each subtype of FTD: 1) FTD subtype < HCs (structural and functional imaging); 2) FTD subtype < HCs (structural imaging); 3) FTD subtype < HCs (functional imaging: FDG-PET, rsfMRI and task-fMRI experiments), 4) FTD subtype > HCs (structural and functional imaging); 4) FTD subtype > HCs (structural imaging); 5) FTD subtype > HCs (functional imaging). We performed conjunction and contrast analyses to identify overlapping and distinctive brain atrophy and functional alterations between different subtypes of FTD. The ALE score map was statistically tested against a null distribution of randomly distributed findings, with a p<0.05 cluster-level family-wise error (cFWE) correction for multiple comparisons (a cluster-forming p < 0.001 with thresholding 1,000 permutations).

## 3. Results

### 3.1. Convergent regional abnormalities in FTD (all subtypes)

A total number of 144 studies involving 166 experiments with 8,057 participants and yielding 2,037 foci were included in our meta-analysis. The results were summarized in Figure 2 and Table S1.

**Figure 2.**
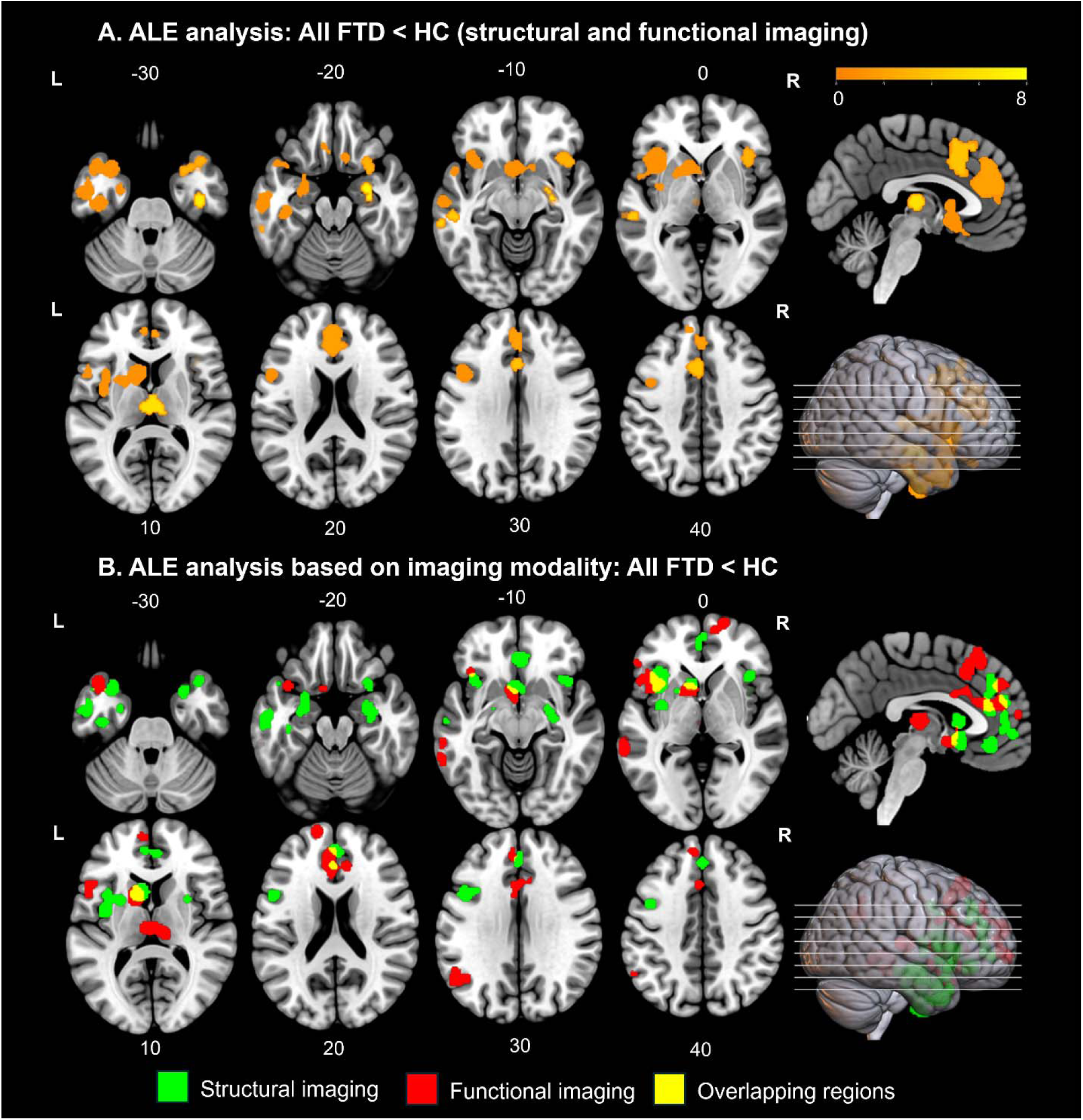
Convergent brain abnormalities in all FTD compared to healthy controls. A) Experiments reporting atrophy/hypoactivation. B) Experiments using structural (green) and functional (red) modalities, with overlapping regions shown in yellow. The coordinates are presented in MNI space. Colour bars represent Z values.

First, we examined the consensus of structural and functional abnormalities by pooling all studies. In the contrast of FTD < HC, we identified eight convergent clusters in the caudate, anterior cingulate cortex (ACC), insula, inferior frontal gyrus (IFG), superior temporal gyrus (STG), middle temporal gyrus (MTG), inferior temporal gyrus (ITG), cingulate cortex (CC), superior frontal gyrus (SFG), thalamus, parahippocampal gyrus (PhG), amygdala, and hippocampus (Fig. 2A).

Next, we performed separate ALE analyses for each imaging modality. The ALE analysis of the structural imaging data (foci = 1390, experiments = 118, subjects = 6179) revealed nine significant clusters in the STG, MTG, PhG, insula, caudate, IFG, and ACC (Fig. 2B). The results from functional imaging (foci = 647, experiments = 48, subjects = 1878) showed ten significant clusters in the ACC, SFG, medial frontal gyrus (MedFG), IFG, insula, thalamus, caudate, STG, and MTG (Fig. 2B). There were no significant clusters identified in the FTD > HC comparisons.

### 3.2. Regional abnormalities in subtypes of FTD

To identify convergent brain abnormalities for the subtypes of FTD, we conducted separate ALE analyses for each subtype (Fig. 3 and Table S2-5). No significant clusters were found in each subtype of FTD > HC comparisons.

**Figure 3.**
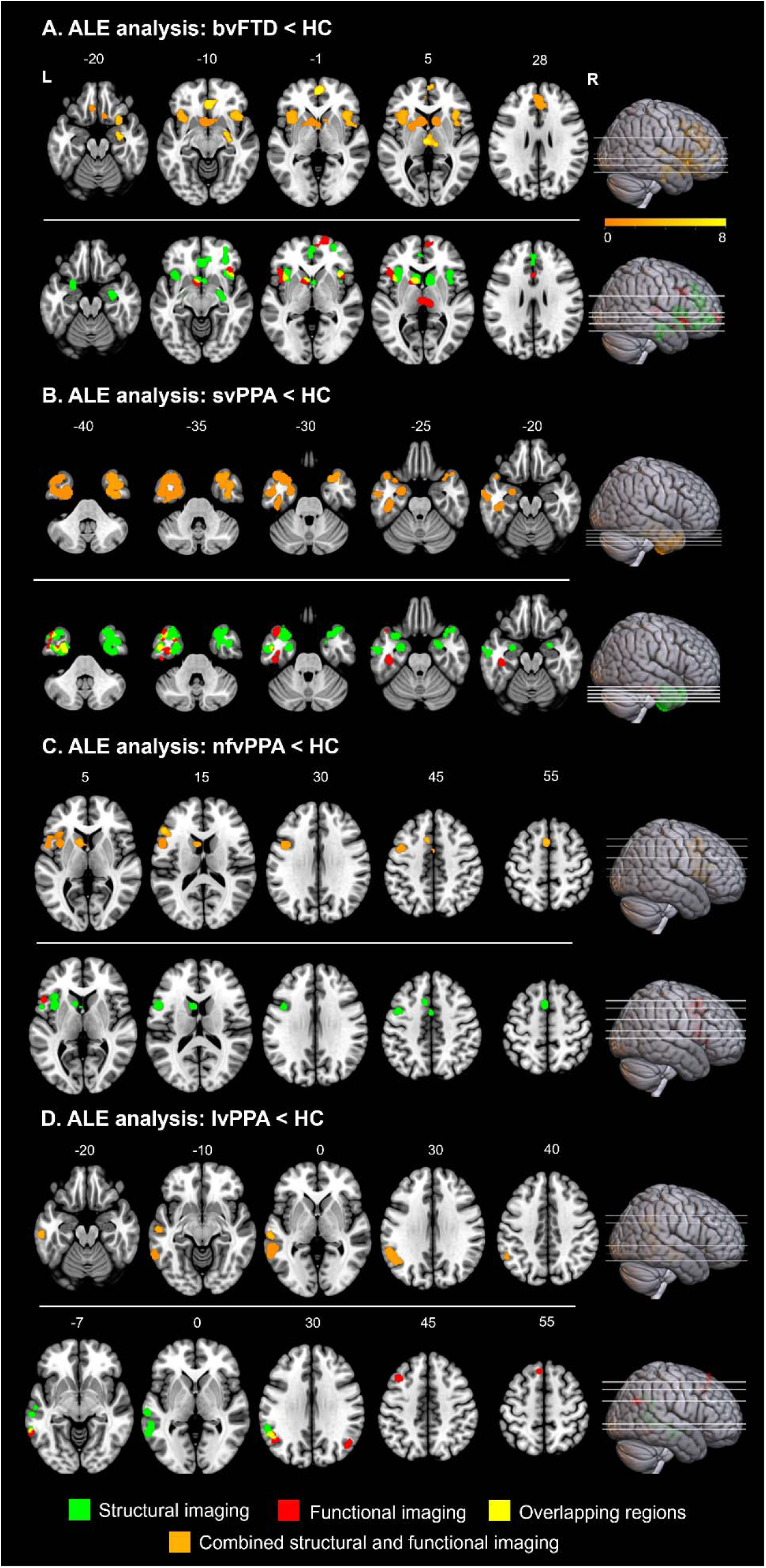
Convergent brain abnormalities in the subtypes of FTD compared to healthy controls. A) The results of ALE analysis in the contrast of bvFTD < HC. B) The results of ALE analysis in the contrast of svPPA/SD < HC. C) The results of ALE analysis in the contrast of nfvPPA < HC. D) The results of ALE analysis in the contrast of lvPPA < HC. Experiments using structural (green) and functional (red) modalities, with overlapping regions shown in yellow. The coordinates are presented in MNI space. Colour bars represent Z values.

For the bvFTD < HC contrast (foci = 1190, experiments = 94, subjects = 4508), nine clusters were identified, including the caudate, putamen, MedFG, thalamus, ACC, cingulate cortex (CC), insula, IFG, STG, amygdala, hippocampus, and fusiform gyrus (FG) (Fig. 3A, top). The structural imaging comparison of bvFTD < HC (foci = 831, experiments = 68, subjects = 3490) revealed ten significant clusters, comprising the MedFG, ACC, insula, amygdala, hippocampus, FG, IFG, caudate, and PhG (Fig. 3A, bottom). For the functional imaging comparison of bvFTD < HC (foci = 370, experiments = 26, subjects = 1018), six significant clusters were found in the caudate, thalamus, insula, MedFG, and CC (Fig. 3A, bottom). The results are summarized in Table S2.

In the contrast of svPPA/SD < HC (foci = 349, experiments = 41, subjects = 1818), we found two clusters in bilateral anterior temporal lobes (ATL) covering the STG, MTG, and ITG (Fig. 3B, top). The structural imaging comparison of svPPA/SD < HC (foci = 219, experiments = 29, subjects = 1422) revealed four clusters in the STG, MTG, ITG, and PhG, while the functional imaging comparison (foci = 130, experiments = 12, subjects = 396) showed a cluster in the left ATL (Fig. 3B, bottom). The results are summarized in Table S3.

In nfvPPA, the contrast of nfvPPA < HC (foci = 228, experiments = 17, subjects = 855) revealed five clusters in the IFG, insular, precentral gyrus, caudate, MedFG, CC, and MFG (Fig. 3C, top). The structural imaging comparison of nfvPPA < HC (foci = 183, experiments = 12, subjects = 678) identified five clusters in the insula, IFG, precentral gyrus, MedFG, CC, SFG, and caudate. The functional imaging comparison (foci = 45, experiments = 5, subjects = 177) revealed clusters only in the IFG and precentral gyrus (Fig. 3C, bottom). The results are summarized in Table S4.

In lvPPA, the contrast of lvPPA < HC (foci = 259, experiments = 14, subjects = 876) identified four clusters in the left STG, MTG, ITG, supramarginal gyrus, and inferior parietal lobe (IPL) (Fig. 3D, top). The subsequent structural imaging comparison (foci = 157, experiments = 9, subjects = 589) revealed three clusters including the STG, MTG, ITG, supramarginal gyrus, and inferior parietal lobe in the left hemisphere (Fig. 3D, bottom). Functional imaging analysis (foci = 102, experiments = 5, subjects = 287) demonstrated five clusters in the left temporal lobe and frontal lobe (Fig. 3D, bottom). The results are summarized in Table S5.

### 3.3. Distinctive and overlapping regions of FTD subtypes

We further investigated the distinctive and overlapping patterns of brain abnormalities in FTD subtypes. To identify unique patterns of brain abnormalities associated with each FTD subtype, we conducted contrast analyses between pairs of subtypes. When comparing bvFTD with svPPA/SD, bvFTD exhibited distinct abnormalities in regions such as the caudate, ACC, MedFG, SFG, insula, IFG, and parahippocampal gyrus, whereas svPPA/SD showed focalized abnormalities primarily in the bilateral ATLs (Fig. 4A). In the comparison between bvFTD and lvPPA, similar regions were affected in bvFTD, including the caudate, parahippocampal gyrus, insula, IFG, ACC, MedFG, and putamen, while lvPPA demonstrated distinctive abnormalities in the left posterior STG, MTG, ITG, supramarginal gyrus, and IPL (Fig. 4B). In the analysis between bvFTD and nfvPPA, bvFTD was associated with abnormalities in the parahippocampal gyrus, insula, IFG, ACC, and MedFG, whereas nfvPPA showed changes in the left IFG, insula, precentral gyrus, MedFG, MFG, and caudate (Fig. 4C). When comparing svPPA/SD with other PPA subtypes, svPPA/SD exhibited limited abnormalities, primarily in the bilateral ATLs (Fig. 4D-E). In contrast, lvPPA relative to svPPA/SD showed greater left-lateralized abnormalities in the posterior STG, MTG, ITG, supramarginal gyrus, and IPL (Fig. 4D), while nfvPPA, compared to svPPA/SD, showed distinct changes in the left IFG, insula, precentral gyrus, MFG, MedFG, SFG, and CC (Fig. 4E). In the comparison between lvPPA and nfvPPA, lvPPA demonstrated additional abnormalities in the left supramarginal gyrus, IPL, and posterior temporal lobe, whereas nfvPPA exhibited changes in the left MedFG, SFG, MFG, IFG, precentral gyrus, and insula (Fig. 4F). Overall, bvFTD displayed unique brain abnormalities in regions including the bilateral IFG, insula, MedFG, ACC, and caudate when compared with PPAs. svPPA/SD was characterized by distinct abnormalities in the bilateral ATLs relative to other subtypes. lvPPA showed unique changes in the left posterior temporal lobe, supramarginal gyrus, and IPL, while nfvPPA presented distinctive abnormalities in the left IFG, insula, precentral gyrus, MFG, and MedFG. These findings are summarized in Figure 4 and Tables S6-11.

**Figure 4.**
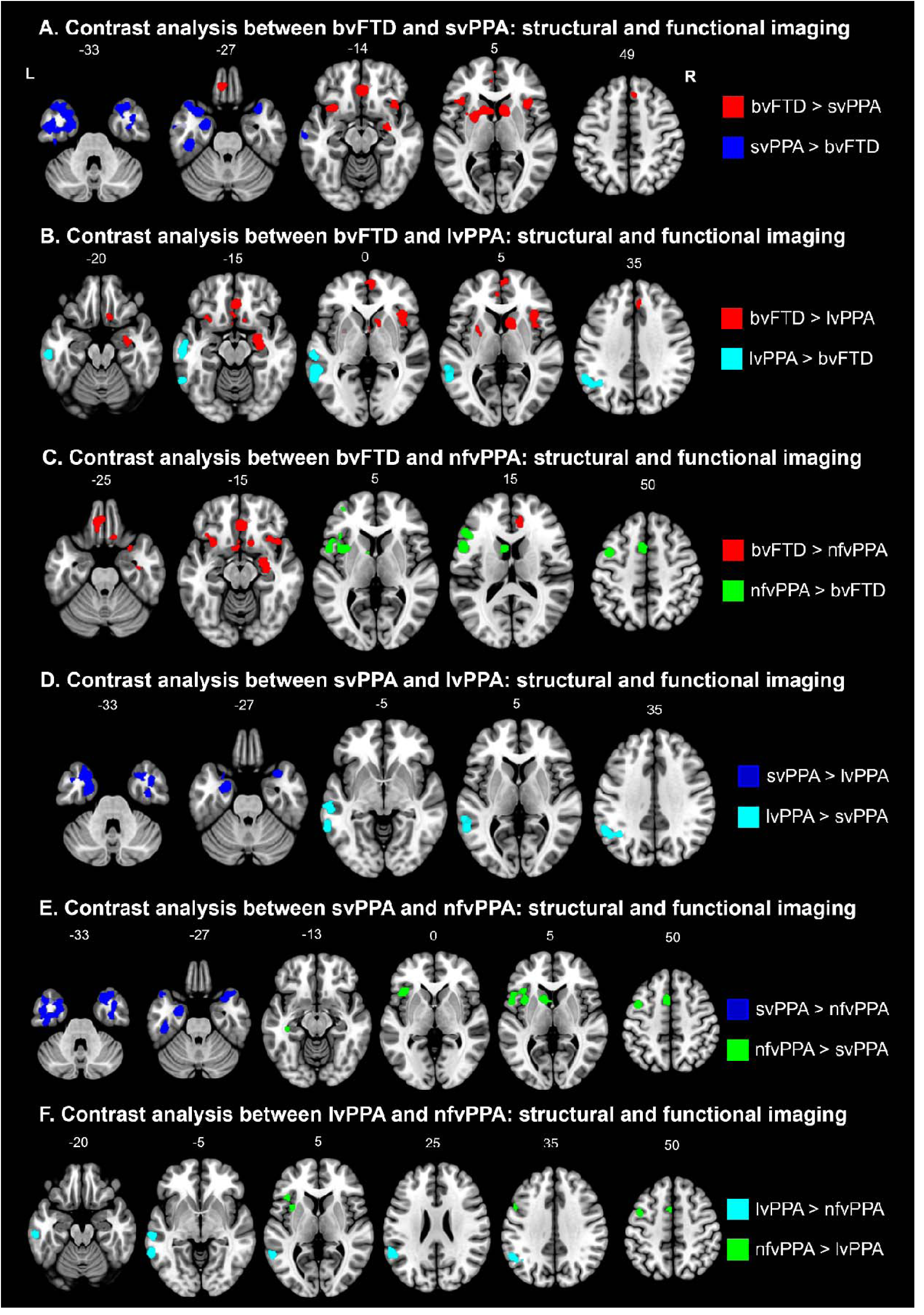
A) The results of contrast analysis between bvFTD and svPPA/SD. B) The results of contrast analysis between bvFTD and lvPPA. C) The results of contrast analysis between bvFTD and nfvPPA. D) The results of contrast analysis between svPPA/SD and lvPPA. E) The results of contrast analysis between svPPA/SD and nfvPPA. F) The results of contrast analysis between lvPPA and nfvPPA.

We overlapped ALE results from each subtype (combined structural and functional imaging) and found overlapping regions between the subtypes of FTD (Fig. 5A). Specifically, bvFTD and svPPA/SD showed an overlap in the right ventral ATL, svPPA/SD and lvPPA overlapped in the left MTG, and bvFTD and nfvPPA overlapped in the left insula, caudate, and ACC. To confirm these findings, we performed conjunction analysis across the FTD subtypes. Conjunction analyses revealed that bvFTD and svPPA/SD overlapped in the bilateral amygdala (in structural imaging only). Additionally, svPPA/SD and lvPPA shared a common region in the left MTG, while bvFTD and nfvPPA shared abnormalities in the left insula, IFG, caudate, and MedFG. Notably, no regions were found to overlap across three or four subtypes. These results were summarized in Figure 5B and Tables S6-11.

**Figure 5.**
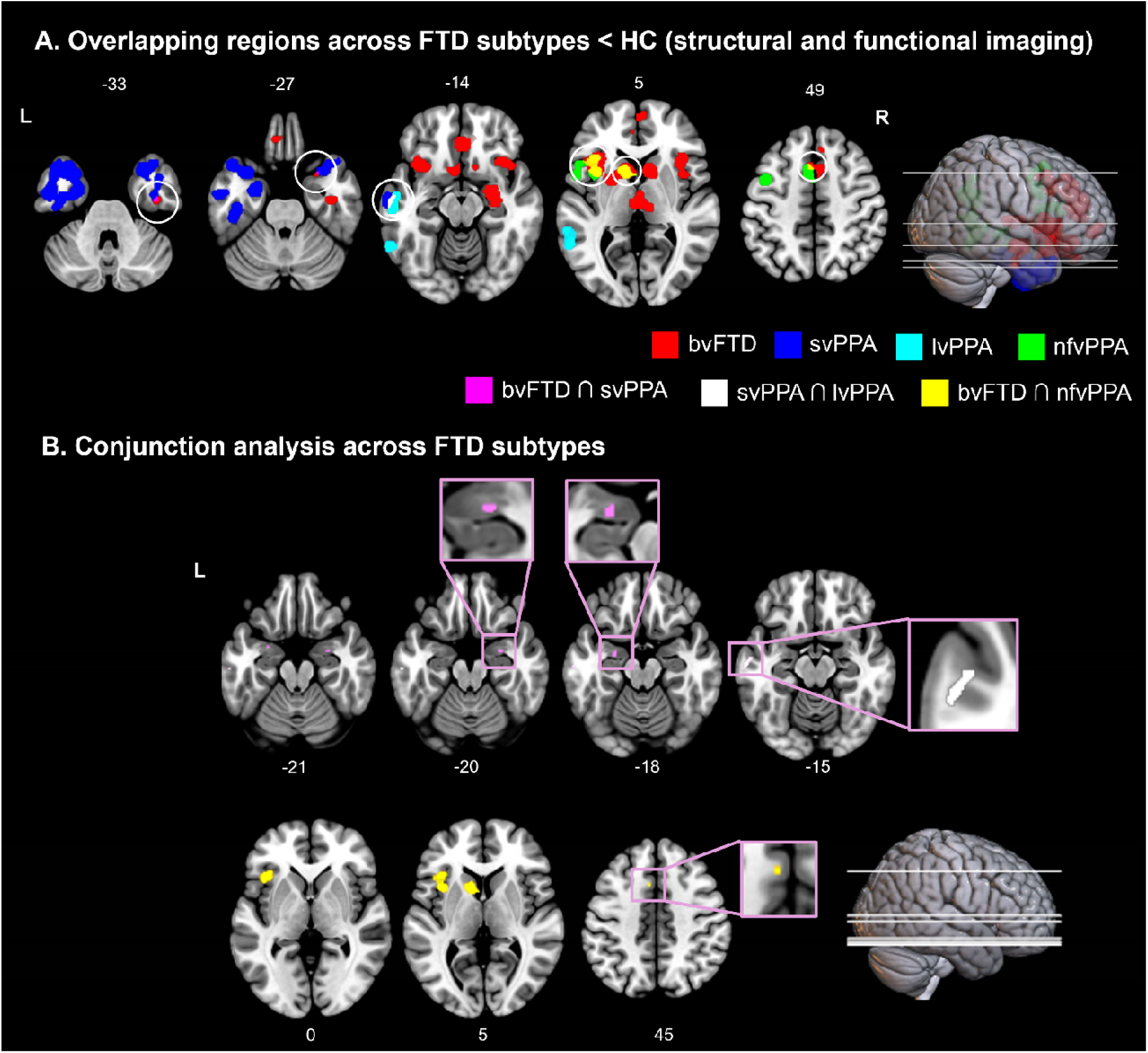
A) Overlapping regions across the FTD subtypes. B) The results of conjunction analysis across the FTD subtypes.

## 4. Discussion

To understand the neural bases of the FTD spectrum, we performed a large-scale CBMA on both structural and functional brain studies across four subtypes of FTD compared to healthy individuals. We analysed neuroimaging data from 8,057 FTD patients, including those with bvFTD, svPPA/SD, nfvPPA, and lvPPA, making this the largest ALE meta-analysis on this topic (N = 1,261) ^50^. Rather than supporting the traditional view of entirely distinct syndromes with non-overlapping neural correlates or a fully continuous spectrum of shared degeneration, our findings reveal that FTD subtypes occupy partially overlapping but distinguishable positions within a multidimensional phenotype geometry, where each subtype is defined by a distinct pattern of atrophy while also sharing specific regions with one or two other subtypes ^11,13,14,16,38,39^. Some regions of degeneration were shared between specific subtypes, reflecting clinical overlaps in symptoms, but no single brain region was common to three or more variants. These findings highlight the systematic variations within each subtype while also acknowledging the overlapping characteristics among different FTD variants. Our findings revealed both common and distinct patterns of brain abnormalities across and within the FTD subtypes, offering new insights that support a transdiagnostic framework of FTD.

### Distinctive patterns of brain abnormalities in FTD syndromes

FTD patients were more likely to exhibit both structural and functional brain abnormalities in various regions of the frontal, temporal and parietal lobes, as well as in the basal ganglia and limbic systems, compared to healthy controls. This widespread degeneration across brain systems aligns with the heterogeneous clinical symptoms and cognitive impairments observed in FTD, including deficits in behaviour, executive function, language, and emotional processing ^1–3^. Our findings are consistent with the established neural characteristics of each FTD subtype and their associated core symptoms.

bvFTD was characterized with degeneration in the salience network (insular, ACC, and CC), limbic system (caudate and thalamus), frontal lobe (SFG, IFG, and MedFG), and medial temporal lobe (amygdala, hippocampus, and parahippocampus). Compared to other FTD subtypes, bvFTD showed distinct abnormalities in the caudate, ACC, insula, medFG, and IFG, and right parahippocampus (Fig.4). Atrophy and hypoactivation/disconnection in these regions are closely associated with the core diagnostic symptoms bvFTD, including impaired empathy, diminished social awareness, and reduced impulse control. Early degeneration of Von Economo neurons (VENs) in the ACC, essential for empathy, social awareness, and self-control, plays a significant role in bvFTD’s social and emotional impairments ^51^. VENs in the anterior insular, another key region in awareness, form a functional network with the striatum and amygdala ^52^. Together, the ACC and insular are central to the salience network, guiding behaviour by assessing the relevance of both internal and external events ^53^. The amygdala, crucial for emotional learning, and behaviour, is also affected early in bvFTD ^54^. Degeneration in the frontal lobe, responsible for self-awareness, cognitive control, emotion regulation, and impulse control, contributes to the disinhibition commonly observed in bvFTD ^20^. Key neural foundations of empathy like the anterior insula, amygdala, ACC, thalamus, and lateral frontal regions ^55^, are also impacted, further explaining bvFTD’s characteristic social and emotional deficits ^56^. The extensive degeneration in these regions results in hallmark bvFTD symptoms such as impaired emotional processing, social cognition deficits, disinhibition, executive dysfunction, and apathy^36^. Additionally, impaired episodic memory in bvFTD is often due to executive dysfunction, which affects the ability to monitor and organize memories ^57^. Similar to Alzheimer’s disease (AD), amnestic bvFTD patients exhibit atrophy or dysfunction in the hippocampus and other paraphippocampus ^58^.

Very different from other subtypes, svPPA/SD showed focalized brain atrophy, hypoactivation/decreased connectivity in the bilateral ATL including STG, MTG, ITG, FG, and paraphippocampal gyrus ^8,59,60^. In svPPA/SD, the defining symptoms include impaired naming and multimodal semantic impairment, often accompanied by specific issues such as impaired object knowledge, surface dyslexia and dysgraphia, intact repetition, and preserved speech fluency ^60^. Clinically, patients with svPPA/SD exhibit relatively fluent speech that lacks specific content. The core symptoms of svPPA/SD directly stem from ATL degeneration, resulting in progressive semantic loss across a wide range of concepts, affecting both expressive and receptive language. This pattern of semantic deficits is evident across modalities, impacting comprehension and production consistently, a hallmark that distinguishes svPPA/SD from other types ^61,62^. Neuroimaging studies have shown a direct correlation between the extent of semantic deficits and the degree of ATL atrophy and hypometabolism, highlighting the role of the ATLs in svPPA/SD’s characteristics ^16,63–65^. These findings support the proposal that the ATLs serve as transmodal transtemporal semantic hubs, integrating information from various modality-specific regions across the cortex ^66,67^. This hub synthesizes multimodal inputs over time and context to create coherent, generalized concepts, essential for comprehensive semantic representation. Such integration is critical for maintaining semantic knowledge across sensory and cognitive domains, explaining the extensive semantic degradation seen in svPPA/SD. ^68^.

nfvPPA exhibited left-lateralized degeneration in the speech production network (SPN), including the insula, IFG, SFG, precentral gyrus, and caudate ^69^. This degeneration pattern results in significant speech production difficulties, with patients displaying slow, effortful speech often marked by grammatical errors and impaired motor planning ^7^. In nfvPPA, the left anterior insula, essential for preparing motor actions in speech, like coordinating vocal tract movements ^70^, also showed degenerations ^32^. A longitudinal study has suggested that atrophy in nfvPPA generally begins in the IFG and progressively impacts other structurally and functionally connected regions within the SPN ^69^. Further supporting this, recent resting-state fMRI findings reveal a substantial loss of connectivity in the left IFG ^71^, contributing to motor speech and syntactic impairments ^72^.

lvPPA was marked by left-lateralized brain abnormalities in the language network, particularly in the posterior temporal and inferior parietal lobes ^73^. Patients with lvPPA typically have slow, hesitant speech with frequent pauses, use simple but grammatically correct sentences, and struggle with naming and repetition tasks ^33^. Damage in these regions disrupts the phonological loop, impairing the ability to retain verbal information, which affects sentence repetition and comprehension ^74,75^. Our results showed that functional imaging meta-analysis revealed degeneration in the left frontal lobe and bilateral inferior parietal regions. A recent study ^73^ suggested that the left posterior temporo-parietal cortex as an epicentre of lvPPA, linked to broader connections in the left frontal and temporal language network, which impacts sentence repetition and naming abilities.

Together, these distinct patterns of degeneration in PPAs suggest how different areas of the brain’s language network contribute to specific language and speech impairments across PPA subtypes. Our findings indicate that frontal lobe damage mainly impacts phonology and speech fluency, dorsal-posterior temporal lobe lesions affect repetition, and lateral/anterior temporal lobe damage impairs language comprehension, especially in semantic processing ^76^.

### Overlapping brain abnormalities between FTD syndromes

Recent research has proposed that the behavioural changes observed in FTD result from disruptions to large-scale brain networks ^77,78^. A neurocognitive model has been proposed to explain these impairments. The controlled social-semantic cognition (CS-SC) model offers a unified frontotemporal framework for understanding social dysfunction across FTD subtypes ^13,16,38^. In this model, social impairments in FTD arises from disruption to two interacting systems: (i) social-semantic knowledge, supported by the ATL, and (ii) social control processes such as selection, evaluation, decision-making, and inhibition, mediated by the frontal cortices, particularly the OFC and medial/lateral prefrontal regions ^38^. This framework helps explain both the semantic and social deficits seen in bvFTD and svPPA/SD, emphasising the interaction between prefrontal and temporal regions in guiding social behaviour.

A related network-based perspective implicates disruptions to broader large-scale systems such as the semantic appraisal network (SAN) ^77^. Within the SAN, the ATL serves as a core hub for representing social-semantic knowledge, while the amygdala and ventromedial OFC contribute to tagging this information with emotional value. Damage to this network can strip social concepts of their meaning and emotional salience, leading to the socioemotional deficits commonly observed in FTD. Recent work also suggests that loss of social-semantic knowledge, particularly following ATL atrophy, may contribute to behavioural disinhibition frequently observed in these patients ^79^.

Consistent with these models, our findings revealed that bvFTD and svPPA/SD share overlapping abnormalities in the ATL and bilateral amygdala. In bvFTD, amygdala atrophy is associated with deficits in decision-making, emotional learning, and behaviour ^54^, as well as difficulties in reading facial emotions ^80^. Similarly, early amygdala atrophy in svPPA contributes to impairments in emotional processing, reflecting the shared frontotemporal changes between these subtypes ^81,82^. As a key node within the SAN ^77^, the amygdala plays a critical role in emotion recognition, integrating socioemotional, interoceptive, and episodic information through connections with the ATL and OFC ^83^, highlighting its significance in the socioemotional dysfunction observed across the FTD spectrum ^84^.

Our finding also revealed the overlapping degeneration between bvFTD and nfvPPA in the left insula, IFG, MedFG, and caudate, key regions supporting controlled selection, inhibition, and monitoring of language output ^85^. Within the CS-SC framework, language impairments in bvFTD may be driven in part by executive control deficits in frontal regions involved in regulating semantic retrieval and social communication ^38^. As a results, bvFTD patients frequently display lexico-semantic deficits, impaired expressive prosody, and difficulties with reading and writing, while motor speech and grammar often remain relatively preserved ^43,86,87^. These language impairments tend to worsen with increasing frontotemporal atrophy and may further exacerbate behavioural disturbances in bvFTD ^43^.

Both svPPA/SD and lvPPA shared abnormalities in the left lateral temporal lobe, particularly in the MTG. Data-driven analyses have identified two subgroups within the lvPPA: one displaying prominent naming and object knowledge impairments associated with lateral temporal hypometabolism ^88^ and atrophy ^89^. Naming impairments in lvPPA has been linked to abnormalities in the left MTG, suggesting its role in the language dysfunction common to both svPPA/SD and lvPPA ^90^. Importantly, lvPPA patients also exhibit semantic control deficits driven by posterior lateral temporal degeneration, contrasting with the more fundamental semantic degradation characteristic of svPPA/SD. A recent study demonstrated that lvPPA patients show variable semantic performance, with impairments emerging on more demanding tasks (e.g., alternative object use, synonym judgement) while performance remains intact on less challenging semantic tests ^39^.

These findings suggest how overlapping patterns of degeneration in FTD subtypes contribute to shared cognitive and language impairments, supporting the view that behavioural changes observed in FTD result from disruptions to large-scale brain networks ^38,77,78^. Rather than conforming to discrete, mutually exclusive diagnostic categories, growing evidence suggests that FTD reflects graded variation across multiple cognitive, behavioural, and language dimensions. Many patients exhibit overlapping features or mixed syndromes that do not fit neatly into a single subtype ^11–13,15^. This emerging multidimensional phenotype geometry suggests that individual FTD syndromes occupy partially overlapping positions within a broader neurocognitive space, shaped by the distribution of neurodegeneration across large-scale brain networks.

### Limitations and future directions

Despite its size and robust results, this study has limitations. First, there was an imbalance in the number of studies per FTD subtype and imaging modality, with a particular lack of functional imaging studies. This limitation may have influenced the detection and definition of functional clusters, potentially leading to underrepresentation of functional network alterations.

Additionally, this study did not control disease stage or severity among participants. FTD syndromes progress differently across subtypes, with some showing more rapid functional and structural decline. Omitting these factors could obscure potential distinctions or overlaps between subtypes that may only emerge at specific disease stages. Future research should prioritize increasing the number of functional imaging studies across FTD subtypes, utilizing advanced imaging techniques like multi-band and multi-echo sequences to capture whole-brain data with high spatial and temporal resolution. These methods enhance sensitivity to subtle functional changes, allowing for better identification of distinct and overlapping networks across subtypes ^91^. Furthermore, longitudinal imaging studies would be beneficial for tracking the progression of abnormalities in both structural and functional networks, providing insight into how degeneration evolves within and across FTD syndromes. By integrating multimodal approaches including structural, functional, and biochemical imaging future studies could yield a more comprehensive understanding of the dynamic and multifaceted nature of FTD.

## Author Contributions

JJ contributed to study design, data collection, analysis and write-up/editing. ZGI contributed to data collection, analysis and write-up. JL and EP contributed to data collection and analysis. AAH contributed to editing. MALR contributed to study design and write-up/editing.

## Acknowledgements

We would like to thank Kamalian A, Khodadadifar T, Saberi A, Masoudi M, Camilleri JA, Eickhoff CR, Zarei M, Pasquini L, Laird AR, Fox PT, Eickhoff SB, and Tahmasian M for kindly sharing unpublished coordinates relating to their published studies for use in this ALE meta-analysis.

## Data availability

The data used in this meta-analysis were extracted from previously published, peer-reviewed neuroimaging studies. As the analysis is based on aggregate coordinate-based data reported in these studies, no individual participant data were collected or generated for this work. Any additional information related to the meta-analytic dataset is available from the corresponding author upon reasonable request.

