## Supplementary Information for "The neural basis of frontotemporal dementia (FTD): insights from ALE meta-analyses of four FTD subtypes encompassing 8,057 patients"

Running title: A meta-analysis of the neural basis of frontotemporal dementia

**Supplementary Tables 1 -11**

**Supplementary Table 1**

Table S1. The ALE results of all FTD < HC

|  | Cluster area | ALE score | Coordinates (centre) | | | Size | p | z |
| --- | --- | --- | --- | --- | --- | --- | --- | --- |
|  |  |  | x | y | z | (mm^3^) |  |  |
| All FTD < HC: *N_foci_* = 2037, *N_experiments_* = 166, *N_subjects_* = 8057 | | | | | | | | |
| 1 | Caudate | 0.11 | -33.8 | 4.1 | -11.5 | 41216 | <.001 | 9.6 |
| 2 | Medial Frontal Gyrus, Anterior Cingulate Cortex | 0.054 | -1.3 | 39.2 | 23.7 | 6792 | <.001 | 5.54 |
| 3 | Insula, Superior Temporal Gyrus, Inferior Frontal Gyrus | 0.058 | 36.2 | 14.6 | -21.5 | 6560 | <.001 | 5.89 |
| 4 | Cingulate Gyrus, Superior Frontal Gyrus, Medial Frontal Gyrus | 0.052 | -2.6 | 16.9 | 43.3 | 3864 | <.001 | 5.36 |
| 5 | Middle Temporal Gyrus, Superior Temporal Gyrus | 0.048 | -58.9 | -30.6 | -7.7 | 1888 | <.001 | 5.04 |
| 6 | Thalamus | 0.061 | 0.9 | -16.7 | 8.2 | 1840 | <.001 | 6.1 |
| 7 | Inferior Temporal Gyrus, Uncus | 0.055 | 37.6 | -10 | -41.1 | 1632 | <.001 | 5.62 |
| 8 | Parahippocampal Gyrus, Amygdala, Hippocampus | 0.051 | 27.3 | -9.7 | -15.6 | 1408 | <.001 | 5.31 |
| FTD < HC (Structural imaging): *N_foci_* = 1390, *N_experiments_* = 118, *N_subjects_* = 6179 | | | | | | | | |
| 1 | Middle Temporal Gyrus, Parahippocampal Gyrus | 0.052 | -38.6 | -6.5 | -29.4 | 13216 | <.001 | 5.86 |
| 2 | Insula, Lentiform Nucleus, Claustrum | 0.078 | -34.7 | 11.3 | 2.8 | 7112 | <.001 | 8.03 |
| 3 | Caudate | 0.078 | -7.4 | 12 | 2.9 | 3784 | <.001 | 8 |
| 4 | Medial Frontal Gyrus, Anterior Cingulate Cortex | 0.043 | -0.3 | 42.5 | 19.1 | 3144 | <.001 | 5.05 |
| 5 | Superior Temporal Gyrus, Uncus | 0.042 | 33 | 10.9 | -33.5 | 2688 | <.001 | 4.99 |
| 6 | Inferior Frontal Gyrus, Precentral Gyrus | 0.047 | -45.8 | 9.4 | 26.8 | 2152 | <.001 | 5.44 |
| 7 | Parahippocampal Gyrus, Amygdala | 0.051 | 27.7 | -9.4 | -16.1 | 2104 | <.001 | 5.84 |
| 8 | Insula | 0.044 | 37.5 | 16.1 | -0.4 | 1784 | <.001 | 5.14 |
| 9 | Anterior Cingulate Cortex | 0.058 | 1 | 35 | -11.3 | 1144 | <.001 | 6.42 |
| FTD < HC (Functional imaging): *N_foci_* = 647, *N_experiments_* = 48, *N_subjects_* = 1878. | | | | | | | | |
| 1 | Cingulate Gyrus, Anterior Cingulate, Medial Frontal Gyrus | 0.033 | -1.8 | 33.4 | 25.3 | 5624 | <.001 | 5.26 |
| 2 | Inferior Frontal Gyrus, Insula, Precentral Gyrus | 0.039 | -45.5 | 17 | 3 | 4856 | <.001 | 5.94 |
| 3 | Caudate | 0.048 | -10.9 | 9.5 | 0.1 | 3824 | <.001 | 6.82 |
| 4 | Thalamus | 0.049 | 2.3 | -19.1 | 7.7 | 2104 | <.001 | 6.89 |
| 5 | Superior Temporal Gyrus | 0.031 | -42.9 | 13.9 | -28.8 | 1856 | <.001 | 4.98 |
| 6 | Middle Temporal Gyrus | 0.028 | -64.3 | -37.6 | -7.5 | 1848 | <.001 | 4.69 |
| 7 | Superior Frontal Gyrus, Medial Frontal Gyrus | 0.027 | -5.2 | 25 | 51.8 | 1616 | <.001 | 4.62 |
| 8 | Superior Frontal Gyrus, Medial Frontal Gyrus | 0.027 | -13.1 | 60.1 | 18.8 | 1192 | <.001 | 4.58 |
| 9 | Middle Temporal Gyrus, Superior Temporal Gyrus | 0.025 | -51.2 | -59.1 | 32.3 | 1064 | <.001 | 4.29 |
| 10 | Medial Frontal Gyrus | 0.029 | 14.4 | 63.1 | -0.2 | 888 | <.001 | 4.81 |

**Supplementary Table 2**

Table S2. The ALE results of bvFTD < HC

|  | Cluster area | ALE score | Coordinates (centre) | | | Size | p | z |
| --- | --- | --- | --- | --- | --- | --- | --- | --- |
|  |  |  | x | y | z | (mm^3^) |  |  |
| All bvFTD < HC: *N_foci_* = 1190, *N_experiments_* = 94, *N_subjects_* = 4508 | | | | | | | | |
| 1 | Caudate | 0.0798 | -4.9 | 12.4 | -0.5 | 10848 | <.001 | 8.52 |
| 2 | Medial Frontal Gyrus, Anterior Cingulate Cortex | 0.0428 | 1.3 | 38.3 | 24.1 | 8032 | <.001 | 5.39 |
| 3 | Insula | 0.0538 | -36 | 17.7 | -3.3 | 4904 | <.001 | 6.4 |
| 4 | Insula | 0.0445 | 38 | 17.5 | -6.1 | 4504 | <.001 | 5.55 |
| 5 | Cingulate Gyrus, Superior Frontal Gyrus | 0.0311 | 2 | 16.3 | 38.8 | 2128 | <.001 | 4.2 |
| 6 | Parahippocampal Gyrus, Amygdala | 0.0464 | 29.5 | -10.1 | -16.9 | 2024 | <.001 | 5.73 |
| 7 | Anterior Cingulate Cortex | 0.0583 | 1.4 | 35 | -11.7 | 1480 | <.001 | 6.78 |
| 8 | Thalamus | 0.0329 | 2.9 | -15.8 | 7.3 | 1312 | <.001 | 4.4 |
| 9 | Medial Frontal Gyrus, Anterior Cingulate Cortex | 0.043 | 1.4 | 55.3 | -1.3 | 1080 | <.001 | 5.41 |
| bvFTD < HC (Structural imaging): *N_foci_* = 831, *N_experiments_* = 68, *N_subjects_* = 3490 | | | | | | | | |
| 1 | Medial Frontal Gyrus, Anterior Cingulate | 0.04 | 0.3 | 42.8 | 18 | 5920 | <.001 | 5.52 |
| 2 | Insula, Inferior Frontal Gyrus, Claustrum | 0.053 | -35 | 17.9 | -2.3 | 3528 | <.001 | 6.7 |
| 3 | Parahippocampal Gyrus, Lentiform Nucleus | 0.046 | 30 | -10.2 | -17.5 | 2688 | <.001 | 6.13 |
| 4 | Superior Frontal Gyrus, Middle Frontal Gyrus | 0.037 | 32.3 | 46.6 | -8.9 | 1944 | <.001 | 5.23 |
| 5 | Insula, Inferior Frontal Gyrus | 0.036 | 38 | 17.6 | -2.3 | 1816 | <.001 | 5.12 |
| 6 | Anterior Cingulate | 0.058 | 1.8 | 35.5 | -11.4 | 1656 | <.001 | 7.2 |
| 7 | Caudate | 0.058 | -9.5 | 12.6 | 7.5 | 1320 | <.001 | 7.18 |
| 8 | Caudate | 0.032 | -1.3 | 10.8 | -6.2 | 1232 | <.001 | 4.7 |
| 9 | Caudate | 0.048 | 10.8 | 11.6 | 8.2 | 1160 | <.001 | 6.24 |
| 10 | Parahippocampal Gyrus, Subcallosal Gyrus | 0.029 | -25.5 | -3.2 | -17.5 | 1080 | <.001 | 4.29 |
| bvFTD < HC (Functional imaging): *N_foci_* = 370, *N_experiments_* = 26, *N_subjects_* = 1018 | | | | | | | | |
| 1 | Caudate | 0.0306 | -10.5 | 9.5 | 0.9 | 3152 | <.001 | 5.6 |
| 2 | Insula, Inferior Frontal Gyrus, Precentral Gyrus | 0.022 | -42 | 17.6 | 1.2 | 1688 | <.001 | 4.48 |
| 3 | Thalamus | 0.0301 | 4.2 | -19 | 7.5 | 1672 | <.001 | 5.53 |
| 4 | Medial Frontal Gyrus | 0.0287 | 15.4 | 64.2 | -0.7 | 1056 | <.001 | 5.36 |
| 5 | Cingulate Gyrus | 0.0198 | 6.5 | 18 | 33.9 | 1008 | <.001 | 4.18 |
| 6 | Insula | 0.0236 | 39.8 | 20.5 | -6.3 | 952 | <.001 | 4.7 |

**Supplementary Table 3**

Table S3. The ALE results of svPPA < HC

|  | Cluster area | ALE score | Coordinates (centre) | | | Size | p | z |
| --- | --- | --- | --- | --- | --- | --- | --- | --- |
|  |  |  | x | y | z | (mm^3^) |  |  |
| All svPPA < HC: *N_foci_* = 349, *N_experiments_* = 41, *N_subjects_* = 1818 | | | | | | | | |
| 1 | Superior Temporal Gyrus, Middle Temporal Gyrus, Inferior Temporal Gyrus | 0.0529 | -39.2 | -3.9 | -32.9 | 22792 | <.001 | 7.89 |
| 2 | Superior Temporal Gyrus, Middle Temporal Gyrus, Inferior Temporal Gyrus | 0.0346 | 35.8 | 4 | -38.5 | 8480 | <.001 | 5.98 |
| svPPA < HC (Structural imaging): *N_foci_* = 219, *N_experiments_* = 29, *N_subjects_* = 1422 | | | | | | | | |
| 1 | Inferior Temporal Gyrus, Uncus, Middle Temporal Gyrus, Parahippocampal Gyrus | 0.043 | -36 | -6.9 | -35.6 | 10064 | <.001 | 7.25 |
| 2 | Superior Temporal Gyrus, Uncus | 0.032 | 35 | 4.4 | -38.6 | 7864 | <.001 | 6.02 |
| 3 | Superior Temporal Gyrus | 0.03 | -35 | 11.8 | -33.2 | 3480 | <.001 | 5.82 |
| 4 | Parahippocampal Gyrus, Uncus | 0.021 | 24.8 | -1.1 | -24.1 | 760 | <.001 | 4.6 |
| svPPA < HC (Functional imaging): *N_foci_* = 130, *N_experiments_* = 12, *N_subjects_* = 396 | | | | | | | | |
| 1 | Superior Temporal Gyrus, Middle Temporal Gyrus, Inferior Temporal Gyrus | 0.022 | -41.5 | -5.8 | -32.8 | 6536 | <.001 | 5.22 |

**Supplementary Table 4**

Table S4. The ALE results of nfvPPA < HC

|  | Cluster area | ALE score | Coordinates (centre) | | | Size | p | z |
| --- | --- | --- | --- | --- | --- | --- | --- | --- |
|  |  |  | x | y | z | (mm^3^) |  |  |
| All nfvPPA < HC: *N_foci_* = 228, *N_experiments_* = 17, *N_subjects_* = 855 | | | | | | | | |
| 1 | Inferior Frontal Gyrus, Insula | 0.0384 | -46.8 | 13.1 | 8.7 | 4824 | <.001 | 6.85 |
| 2 | Precentral gyurs, Middle Frontal Gyrus | 0.0298 | -44.3 | 5.9 | 38.4 | 3016 | <.001 | 5.79 |
| 3 | Caudate | 0.023 | -10 | 10.3 | 9.2 | 1232 | <.001 | 4.85 |
| 4 | Medial Frontal Gyrus, Cingulate Cortex | 0.0201 | -5.1 | 11.2 | 49.8 | 1216 | <.001 | 4.47 |
| 5 | Middle Frontal Gyrus, Inferior Frontal Gyrus | 0.0202 | -49.3 | 24.6 | 18 | 920 | <.001 | 4.48 |
| nfvPPA < HC (Structural imaging): *N_foci_* = 183, *N_experiments_* =12, *N_subjects_* = 678 | | | | | | | | |
| 1 | Insula, Inferior Frontal Gyrus | 0.031 | -37.4 | 16 | 3.5 | 3292 | <.001 | 6.12 |
| 2 | Precentral Gyrus, Middle Frontal Gyrus | 0.021 | -43.6 | 4.5 | 40 | 1992 | <.001 | 4.8 |
| 3 | Medial Frontal Gyrus, Cingulate Gyrus, Superior Frontal Gyrus | 0.02 | -4.8 | 10.5 | 49.5 | 1568 | <.001 | 4.66 |
| 4 | Inferior Frontal Gyrus | 0.031 | -53.5 | 9.6 | 15.4 | 1352 | <.001 | 6.15 |
| 5 | Caudate | 0.019 | -8.6 | 10.2 | 10.8 | 920 | <.001 | 4.53 |
| nfvPPA < HC (Functional imaging): *N_foci_* = 45, *N_experiments_* = 5, *N_subjects_* = 177 | | | | | | | | |
| 1 | Inferior Frontal Gyrus, Precentral Gyrus | 0.018 | -51.4 | 17.9 | 6.1 | 584 | <.001 | 5.27 |

**Supplementary Table 5**

Table S5. The ALE results of lvPPA < HC

|  | Cluster area | ALE score | Coordinates (centre) | | | Size | p | z |
| --- | --- | --- | --- | --- | --- | --- | --- | --- |
|  |  |  | x | y | z | (mm^3^) |  |  |
| All lvPPA < HC: *N_foci_* = 259, *N_experiments_* = 14, *N_subjects_* = 876 | | | | | | | | |
| 1 | Middle Temporal Gyrus, Inferior Temporal Gyrus, Superior Temporal gyrus | 0.0286 | -60.5 | -48.5 | -0.9 | 4664 | <.001 | 5.49 |
| 2 | Supramarginal Gryus, Inferior Parietal Gyrus | 0.0337 | -53.3 | -52.9 | 30.4 | 3928 | <.001 | 6.11 |
| 3 | Middle Temporal Gyrus, Inferior Temporal Gyrus | 0.0237 | -59.2 | -19.5 | -15.3 | 1336 | <.001 | 4.82 |
| 4 | Middle Temporal gyrus, Superior Temporal Gyrus | 0.026 | -61.7 | -26.6 | -3.7 | 840 | <.001 | 5.13 |
| lvPPA < HC (Structural imaging): *N_foci_* = 157, *N_experiments_* = 9, *N_subjects_* = 589 | | | | | | | | |
| 1 | Middle Temporal Gyrus, Superior Temporal Gyrus, Supramarginal Gyrus, Inferior Parietal Lobule | 0.02 | -58.1 | -48.8 | 14.4 | 5176 | <.001 | 4.71 |
| 2 | Middle Temporal Gyrus | 0.018 | -58.5 | -17 | -14.2 | 1352 | <.001 | 4.26 |
| 3 | Superior Temporal Gyrus, Middle Temporal Gyrus | 0.019 | -62.6 | -25.3 | -2.2 | 776 | <.001 | 4.46 |
| lvPPA < HC (Functional imaging): *N_foci_* = 102, *N_experiments_* = 5, *N_subjects_* = 287 | | | | | | | | |
| 1 | Middle Temporal Gyrus, Superior Temporal Gyrus | 0.017 | -51 | -56.5 | 30 | 1120 | <.001 | 4.64 |
| 2 | Inferior Temporal Gyrus, Middle Temporal Gyrus | 0.017 | -63.3 | -50.5 | -8.8 | 584 | <.001 | 4.56 |
| 3 | Superior Frontal Gyrus | 0.014 | -6.2 | 29.3 | 57.3 | 552 | <.001 | 4.08 |
| 4 | Middle Temporal Gyrus | 0.017 | 47.5 | -66.2 | 29.2 | 440 | <.001 | 4.55 |
| 5 | Superior Frontal Gyrus, Middle Frontal Gyrus | 0.017 | -38.4 | 20.9 | 45.5 | 440 | <.001 | 4.71 |

**Supplementary Table 6**

Table S6. The ALE results of the comparison of bvFTD < HC and svPPA < HC

|  | Cluster area | Coordinates (centre) | | | Size | p | z |
| --- | --- | --- | --- | --- | --- | --- | --- |
|  |  | x | y | z | (mm^3^) |  |  |
| bvFTD > svPPA (Structural and Functional imaging) | | | | | | | |
| 1 | Caudate | -3.9 | 10.1 | 4.6 | 5872 | <.001 | 3.89 |
| 2 | Anterior Cingulate Cortex, Medial Frontal Gyrus, Superior Frontal Gyrus | 3.6 | 36.5 | 27 | 4288 | <.001 | 3.72 |
| 3 | Insula, Inferior Frontal Gyrus | 39.8 | 20.1 | -5 | 1952 | <.006 | 2.51 |
| 4 | Insula, Inferior Frontal Gyrus | -36.7 | 18.9 | -6.2 | 1496 | <.002 | 2.85 |
| 5 | Anterior Cingulate Cortex | 1.7 | 35.7 | -11.7 | 1464 | <.001 | 3.89 |
| 6 | Cingulate Gyrus, Medial Frontal Gyrus | 5.4 | 17.8 | 34.7 | 808 | <.002 | 2.89 |
| 7 | Anterior Cingulate Cortex, Medial Frontal Gyrus | 1.2 | 54.1 | -1.8 | 664 | <.005 | 2.64 |
| 8 | Medial Frontal Gyrus | -6.1 | 38.5 | -23.5 | 632 | <.001 | 3.89 |
| 9 | Lentiform Nucleus, Parahippocampal Gyrus | 28.6 | -7.2 | -12.9 | 272 | <.008 | 2.38 |
| 10 | Insula | 48.6 | 6.9 | -1.1 | 128 | <.008 | 2.46 |
| svPPA > bvFTD (Structural and Functional imaging) | | | | | | | |
| 1 | Superior Temporal Gyrus, Middle Temporal Gyrus, Inferior Temporal Gyrus, Parahippocampal Gyrus, Fusiform Gyrus | -38.2 | -1.3 | -34.1 | 16504 | <.001 | 3.54 |
| 2 | Superior Temporal Gyrus, Middle Temporal Gyrus, Inferior Temporal Gyrus | 35.3 | 3.2 | -39.2 | 6440 | <.001 | 3.43 |
| 3 | Fusiform Gyrus, Parahippocampal Gyrus, Inferior Temporal Gyrus | -44.6 | -27.8 | -24.6 | 1600 | <.001 | 3.89 |
| 4 | Amygdala | 26.2 | -2.3 | -23 | 88 | <.025 | 1.96 |
| Conjunction bvFTD & svPPA (Structural imaging) | | | | | | | |
| 1 | Amygdala | -24.8 | -6.3 | -18 | 40 | <.014 |  |
| 2 | Amygdala | 26.9 | -4 | -20 | 16 | <.014 |  |
| 3 | Amygdala | -26 | 0 | -22 | 8 | <.012 |  |

**Supplementary Table 7**

Table S7. The ALE results of the comparison of bvFTD < HC and lvPPA < HC

|  | Cluster area | Coordinates (centre) | | | Size | p | z |
| --- | --- | --- | --- | --- | --- | --- | --- |
|  |  | x | y | z | (mm^3^) |  |  |
| bvFTD > lvPPA (Strucutral and Functional imaging) | | | | | | | |
| 1 | Caudate, Anterior Cingulate Cortex | 7.9 | 11.7 | -2.1 | 3856 | <.001 | 3.29 |
| 2 | Parahippocampal Gyrus, Amygdala | 27.8 | -9.3 | -15.8 | 1624 | <.001 | 2.44 |
| 3 | Insula, Inferior Frontal Gyrus | 39.4 | 19 | 2.5 | 1544 | <.001 | 2.97 |
| 4 | Anterior Cingulate Cortex | 3.3 | 33.9 | -12.8 | 920 | <.002 | 2.86 |
| 5 | Anterior Cingulate Cortex | 7 | 41.6 | 12 | 920 | <.005 | 2.55 |
| 6 | Medial Frontal Gyrus | 3.6 | 57.6 | 1 | 480 | <.004 | 2.64 |
| 7 | Claustrum | -31.1 | 14.2 | -11.5 | 232 | <.015 | 2.18 |
| 8 | Anterior Cingulate Cortex | -5.6 | 45.7 | 9.2 | 208 | <.011 | 2.28 |
| 9 | Medial Frontal Gyrus | -27.2 | 4.5 | 4.5 | 160 | <.03 | 1.87 |
| 10 | Putamen | 26 | 8 | -24.1 | 112 | <.03 | 1.88 |
| lvPPA > bvFTD (Strucutral and Functional imaging) | | | | | | | |
| 1 | Middle Temporal Gyrus, Inferior Temporal Gyrus, Superior Temporal Gyrus, Fusiform Gyrus | -60.4 | -48.5 | -0.6 | 4664 | <.001 | 3.89 |
| 2 | Superior Temporal Gyrus, Supramarginal Gyrus, Inferior Parietal Lobule, Middle Temporal Gyrus | -53.6 | -52.7 | 30.1 | 3928 | <.001 | 3.89 |
| 3 | Middle Temporal Gyrus, Inferior Temporal Gyrus | -59.1 | -19.5 | -15 | 1336 | <.001 | 3.89 |
| 4 | Middle Temporal Gyrus, Superior Temporal Gyrus | -62.1 | -26.2 | -3.7 | 824 | <.001 | 3.89 |
| Conjunction bvFTD & lvPPA | | | | | | | |
| - | - | - | - | - | - | - | - |

**Supplementary Table 8**

Table S8. The ALE results of the comparison of bvFTD < HC and nfvPPA < HC

| bvFTD > nfvPPA (Strucutral and Functional imaging) | | | | | | | |
| --- | --- | --- | --- | --- | --- | --- | --- |
| 1 | Parahippocampal Gyrus, Amygdala | 29.8 | -10 | -17.3 | 2024 | <.001 | 2.93 |
| 2 | Insula, Inferior Frontal Gyrus | 34.5 | 13.9 | -15 | 1592 | <.001 | 3.19 |
| 3 | Anterior Cingulate Cortex | 1.7 | 35.1 | -11.9 | 1480 | <.001 | 3.72 |
| 4 | Medial Frontal Gyrus, Anterior Cingulate Cortex | -7.1 | 33.7 | -22.1 | 1136 | <.002 | 2.85 |
| 5 | Caudate | 6.8 | 15.3 | -15.7 | 1000 | <.001 | 3.89 |
| 6 | Anterior Cingulate Cortex | 9.8 | 38.7 | 14.3 | 584 | <.009 | 2.36 |
| 7 | Claustrum, Inferior Frontal Gyrus, Insula | -31.8 | 16.2 | -12.7 | 392 | <.018 | 2.09 |
| 8 | Cingulate Gyrus | 9 | 19.9 | 37.2 | 192 | <.025 | 1.98 |
| 9 | Cingulate Gyrus, Medial Frontal Gyrus | 9.8 | 33.5 | 29 | 64 | <.025 | 1.98 |
| 10 | Cingulate Gyrus | 6 | 40 | 24 | 24 | <.038 | 1.78 |
| nfvPPA > bvFTD (Strucutral and Functional imaging) | | | | | | | |
| 1 | Inferior Frontal Gyrus, Insula, Precentral Gyrus | -51.3 | 12.2 | 10.9 | 3512 | <.001 | 3.89 |
| 2 | Precentral Gyrus, Middle Frontal Gyrus, Inferior Frontal Gyrus | -44.3 | 5.3 | 39.4 | 2928 | <.001 | 3.89 |
| 3 | Medial Frontal Gyrus, Cingulate Gyrus, | -5 | 10.7 | 50.6 | 1112 | <.001 | 3.89 |
| 4 | Middle Frontal Gyrus, Inferior Frontal Gyrus | -49.3 | 24.5 | 18.2 | 920 | <.001 | 3.89 |
| 5 | Insula | -36.7 | 8.8 | 6.6 | 816 | <.001 | 3.24 |
| 6 | Caudate | -7.5 | 7.5 | 12.5 | 384 | <.007 | 2.46 |
| Conjunction bvFTD & nfvPPA | | | | | | | |
| 1 | Insula, Inferior Frontal Gyrus | -37.8 | 18 | 2.4 | 1488 | <.03 |  |
| 2 | Caudate | -10.8 | 10.7 | 8.4 | 1016 | <.03 |  |
| 3 | Medial Frontal Gyrus | -6 | 14 | 44 | 8 | <.012 |  |

**Supplementary Table 9**

Table S9. The ALE results of the comparison of svPPA < HC and lvPPA < HC

|  | Cluster area | Coordinates (centre) | | | Size | p | z |
| --- | --- | --- | --- | --- | --- | --- | --- |
|  |  | x | y | z | (mm^3^) |  |  |
| svPPA > lvPPA (Strucutral and Functional imaging) | | | | | | | |
| 1 | Uncus, Superior Temporal Gyrus, Middle Temporal Gyrus, Inferior Temporal Gyrus, Parahippocampal Gyrus | -29.7 | -1.5 | -37.7 | 8024 | <.001 | 3.89 |
| 2 | Superior Temporal Gyrus, Middle Temporal Gyrus, Inferior Temporal Gyrus | 36.3 | 5.8 | -36.2 | 2872 | <.001 | 3.19 |
| 3 | Uncus, Inferior Temporal Gyrus | 40.5 | -11.9 | -34.8 | 248 | <.015 | 2.18 |
| 4 | Inferior Temporal Gyrus | -64 | -8 | -24 | 24 | <.048 | 1.67 |
| 5 | Uncus | 28 | -4 | -44 | 8 | <.047 | 1.68 |
| lvPPA > svPPA (Strucutral and Functional imaging) | | | | | | | |
| 1 | Middle Temporal Gyrus, Inferior Temporal Gyrus, Superior Temporal Gyrus | -60.2 | -48.3 | -0.2 | 4568 | <.001 | 3.89 |
| 2 | Supramarginal Gyrus, Inferior Parietal Lobule | -54.3 | -52 | 30 | 3896 | <.001 | 3.89 |
| 3 | Middle Temporal Gyrus | -58.8 | -21.7 | -15.4 | 1000 | <.001 | 3.16 |
| 4 | Middle Temporal Gyrus | -61.7 | -26.2 | -3.5 | 832 | <.001 | 3.12 |
| Conjunction svPPA & lvPPA | | | | | | | |
| 1 | Middle Temporal Gyrus | -60.2 | -15.1 | -14.5 | 216 | <.017 |  |

**Supplementary Table 10**

Table S10. The ALE results of the comparison of svPPA < HC and nfvPPA < HC

|  | Cluster area | Coordinates (centre) | | | Size | p | z |
| --- | --- | --- | --- | --- | --- | --- | --- |
|  |  | x | y | z | (mm^3^) |  |  |
| svPPA > nfvPPA (Strucutral and Functional imaging) | | | | | | | |
| 1 | Uncus, Superior Temporal Gyrus, Middle Temporal Gyrus, Inferior Temporal Gyrus, Parahippocampal Gyrus, Fusiform Gyrus | -36.5 | -4.8 | -37.1 | 13904 | <.001 | 3.89 |
| 2 | Superior Temporal Gyrus, Middle Temporal Gyrus, Inferior Temporal Gyrus | 36 | 3.6 | -38.4 | 8480 | <.001 | 3.89 |
| nfvPPA > svPPA (Strucutral and Functional imaging) | | | | | | | |
| 1 | Inferior Frontal Gyrus, Insula, Precentral Gyrus | -48.3 | 12.9 | 9.4 | 4696 | <.001 | 3.89 |
| 2 | Precentral Gyrus, Middle Frontal Gyrus, Inferior Frontal Gyrus | -44.4 | 5.9 | 38.2 | 3016 | <.001 | 3.89 |
| 3 | Medial Frontal Gyrus, Cingulate Gyrus, Superior Frontal Gyrus | -5.1 | 10.5 | 49.6 | 1096 | <.001 | 3.43 |
| 4 | Caudate | -10.1 | 11.1 | 8.6 | 1040 | <.001 | 3.29 |
| 5 | Middle Frontal Gyrus, Inferior Frontal Gyrus | -49.4 | 24.6 | 18 | 920 | <.001 | 3.89 |
| Conjunction svPPA & nfvPPA | | | | | | | |
| - | - | - | - | - | - | - | - |

**Supplementary Table 11**

Table S11. The ALE results of the comparison of lvPPA < HC and nfvPPA < HC

|  | Cluster area | Coordinates (centre) | | | Size | p | z |
| --- | --- | --- | --- | --- | --- | --- | --- |
|  |  | x | y | z | (mm^3^) |  |  |
| lvPPA > nfvPPA (Strucutral and Functional imaging) | | | | | | | |
| 1 | Supramarginal Gyrus, Superior Temporal Gyrus, Inferior Parietal Lobule | -55.5 | -51 | 29.4 | 3360 | <.001 | 3.89 |
| 2 | Middle Temporal Gyrus, Inferior Temporal Gyrus, Superior Temporal Gyrus | -60.4 | -51.2 | -0.6 | 3288 | <.001 | 3.54 |
| 3 | Middle Temporal Gyrus | -62.7 | -26.1 | -3.8 | 752 | <.001 | 3.72 |
| 4 | Middle Temporal Gyrus | -60.5 | -23.9 | -17.2 | 600 | <.001 | 3.16 |
| nfvPPA > lvPPA (Strucutral and Functional imaging) | | | | | | | |
| 1 | Medial Frontal Gyrus, Superior Frontal Gyrus | -3.6 | 9.1 | 52.7 | 592 | <.02 | 2.05 |
| 2 | Inferior Frontal Gyrus | -43.9 | 3.1 | 48 | 544 | <.02 | 2.46 |
| 3 | Middle Frontal Gyrus, Precentral Gyrus | -43.9 | 3.1 | 48 | 536 | <.025 | 1.97 |
| 4 | Claustrum, Insula | -34 | 9.5 | 7.3 | 320 | <.012 | 2.27 |
| 5 | Middle Frontal Gyrus, Precentral Gyrus | -49.7 | 9.5 | 37.3 | 232 | <.016 | 2.13 |
| 6 | Inferior Frontal Gyrus, Insula | -40.6 | 22.7 | 3.9 | 136 | <.027 | 1.93 |
| Conjunction lvPPA & nfvPPA | | | | | | | |
| - | - | - | - | - | - | - | - |
